# Large-scale crowdsourced radiotherapy segmentations across a variety of cancer anatomic sites: Interobserver expert/non-expert and multi-observer composite tumor and normal tissue delineation annotations from a prospective educational challenge

**DOI:** 10.1101/2022.10.05.22280672

**Authors:** Kareem A. Wahid, Diana Lin, Onur Sahin, Michael Cislo, Benjamin E. Nelms, Renjie He, Mohammed A. Naser, Simon Duke, Michael V. Sherer, John P. Christodouleas, Abdallah S. R. Mohamed, James D. Murphy, Clifton D. Fuller, Erin F. Gillespie

## Abstract

Clinician generated segmentation of tumor and healthy tissue regions of interest (ROIs) on medical images is crucial for radiotherapy. However, interobserver segmentation variability has long been considered a significant detriment to the implementation of high-quality and consistent radiotherapy dose delivery. This has prompted the increasing development of automated segmentation approaches. However, extant segmentation datasets typically only provide segmentations generated by a limited number of annotators with varying, and often unspecified, levels of expertise. In this data descriptor, numerous clinician annotators manually generated segmentations for ROIs on computed tomography images across a variety of cancer sites (breast, sarcoma, head and neck, gynecologic, gastrointestinal; one patient per cancer site) for the Contouring Collaborative for Consensus in Radiation Oncology challenge. In total, over 200 annotators (experts and non-experts) contributed using a standardized annotation platform (ProKnow). Subsequently, we converted data into NIfTI format with standardized nomenclature for ease of use. In addition, we generated consensus segmentations for experts and non-experts using the STAPLE method. These standardized, structured, and easily accessible data are a valuable resource for systematically studying variability in segmentation applications.

## Background & Summary

Since the advent of contemporary radiation delivery techniques for cancer treatment, clinician generated segmentation (also termed contouring or delineation) of target structures (e.g., primary tumors and metastatic lymph nodes) and organs at risk (e.g., healthy tissues whose irradiation could lead to damage and/or side effects) on medical images has become a necessity in the radiotherapy workflow ^1^. These segmentations are typically provided by trained medical professionals, such as radiation oncologists. While segmentations can be performed on any imaging modality that provides sufficient discriminative capabilities to visualize regions of interest (ROIs), the current radiotherapy workflow prioritizes the use of computed tomography (CT) for ROI segmentation due to its ubiquitous nature and use in radiotherapy dose calculations. Subsequently, clinicians spend a large fraction of their time and effort generating ROI segmentations on CT imaging necessary for the radiotherapy workflow.

Interobserver and intraobserver variability are well-documented byproducts of the use of manual human-generated segmentations ^2,3^. While consensus radiotherapy guidelines to ensure ROI segmentation quality have been developed and shown to reduce variability ^4^, these guidelines are not necessarily followed by all practicing clinicians. Therefore, segmentation variability remains a significant concern in maintaining radiotherapy plan quality and consistency. Recent computational improvements in machine learning, particularly deep learning, have prompted the increasing development and deployment of accurate ROI auto-segmentation algorithms to reduce radiotherapy segmentation variability ^5–7^. However, for auto-segmentation algorithms to be clinically useful, their input data (training data) should reflect high-quality “gold-standard” annotations. While research has been performed on the impact of interobserver variability and segmentation quality for auto-segmentation training ^8–11^, it remains unclear how “gold-standard” segmentations should be defined and generated. One common approach, consensus segmentation generation, seeks to crowdsource multiple segmentations from different annotators to generate a high-quality ground-truth segmentation. While multi-observer public medical imaging segmentation datasets exist ^12–17^, there remains a lack of datasets with a large number of annotators for radiotherapy applications.

The Contouring Collaborative for Consensus in Radiation Oncology (C3RO) challenge was developed to engage radiation oncologists across various expertise levels in cloud-based ROI crowdsourced segmentation ^18^. Through this collaboration, a large number of clinicians generated ROI segmentations using CT images from 5 unique radiotherapy cases: breast, sarcoma, head and neck, gynecologic, and gastrointestinal. In this data descriptor, we present the curation and processing of the data from the C3RO challenge. The primary contribution of this dataset is unprecedented large-scale multi-annotator individual and consensus segmentations of various ROIs crucial for radiotherapy planning in an easily accessible and standardized imaging format. These data can be leveraged for exploratory analysis of segmentation quality across a large number of annotators, consensus segmentation experiments, and auto-segmentation model benchmarking. An overview of this data descriptor is shown in **Fig. 1**.

**Figure 1.**
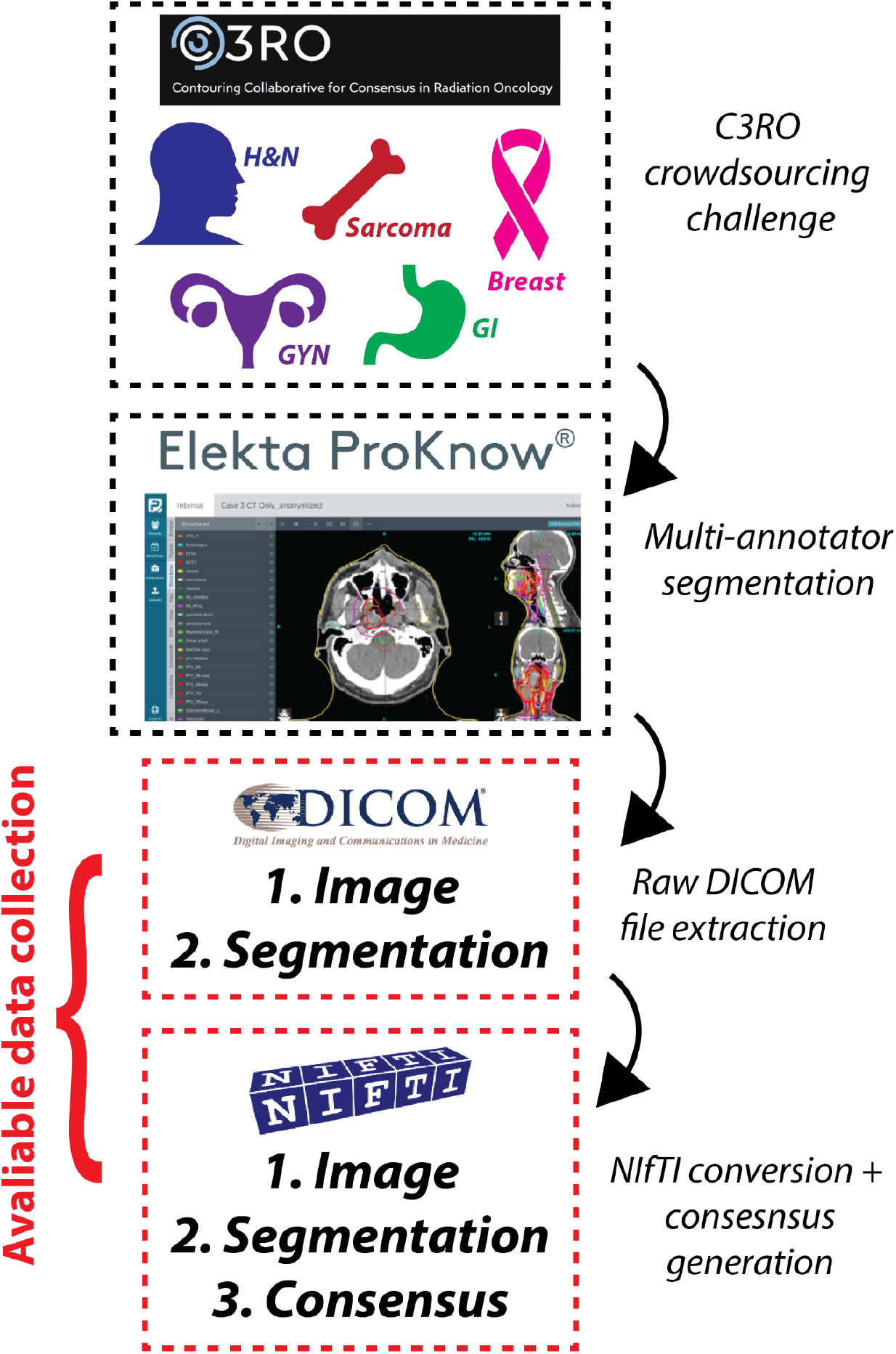
Data descriptor overview. Multi-annotator segmentations were generated for the Contouring Collaborative for Consensus in Radiation Oncology (C3RO) challenge. Imaging and segmentation data were extracted from the ProKnow cloud-based platform in Digital Imaging and Communications in Medicine (DICOM) format, which were then converted to Neuroimaging Informatics Technology Initiative (NIfTI) format for ease of use. Consensus segmentation data were then generated from the available multi-annotator segmentations. The provided data collection contains all original DICOM files along with converted NIfTI files.

## Methods

### Patient population

Five separate patients who had undergone radiotherapy were retrospectively collected from our collaborators at various institutions. Each patient had received a pathologically confirmed diagnosis of cancer of one of the following sites: breast (post-mastectomy intraductal carcinoma), sarcoma (malignant peripheral nerve sheath tumor of the left thigh), head and neck (oropharynx with nodal spread, [H&N]), gynecologic (cervical cancer, [GYN]), and gastrointestinal (anal cancer, [GI]). Clinical characteristics of these patients are shown in **Table 1**. Of note, these five disease sites were included as part of the C3RO challenge due to being among the most common disease sites treated by radiation oncologists; additional disease sites were planned but were not realized due to diminishing community participation in C3RO. Specific patient cases were selected by C3RO collaborators on the basis of being adequate reflections of routine patients a generalist radiation oncologist may see in a typical workflow (i.e., not overly complex). Further details on the study design for C3RO can be found in Lin & Wahid et al. ^19^.

**Table 1.**
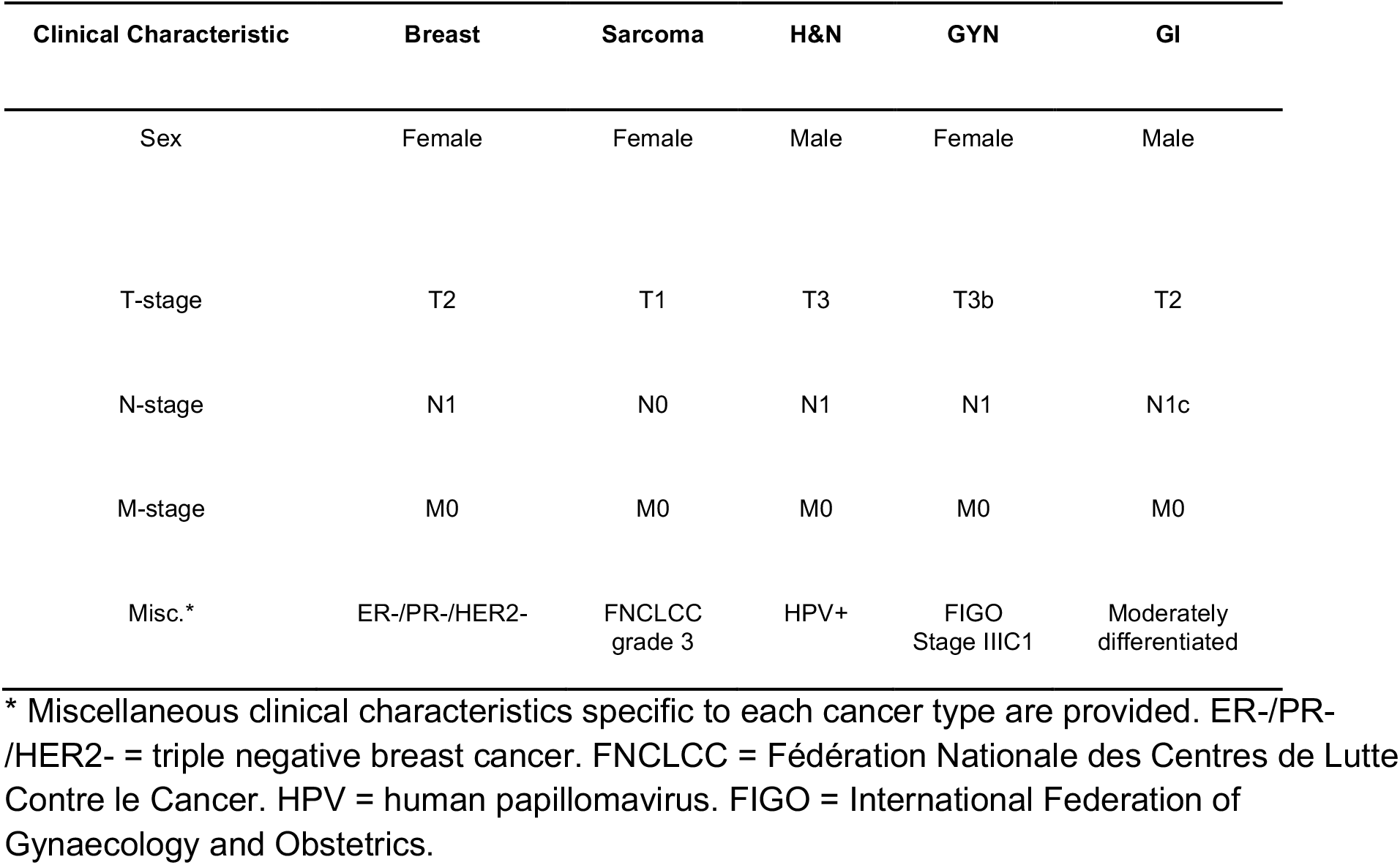
Clinical characteristics for cases included in this data descriptor. Cases included breast, sarcoma, head and neck (H&N), gynecologic (GYN), and gastrointestinal (GI) cancer.

### Imaging protocols

Each patient received a radiotherapy planning CT scan which was exported in Digital Imaging and Communications in Medicine (DICOM) format. CT image acquisition characteristics are shown in **Table 2**. All images were acquired on scanners that were routinely used for radiotherapy planning at their corresponding institutions with appropriate calibration and quality assurance by technical personnel. The sarcoma, H&N, and GI cases received intravenous contrast, the GU case received oral contrast, and the breast case did not receive any contrast. Of note, the H&N case had metal streak artifacts secondary to metallic implants in the upper teeth, which obscured anatomy near the mandible. No other cases contained noticeable image artifacts. Notably, the sarcoma case also received a magnetic resonance imaging (MRI) scan, while the H&N and GI cases received full body positron emission tomography (PET) scans. The sarcoma MRI scan was acquired on a GE Signa HDxt device and corresponded to a post-contrast spin echo T1-weighted image with a slice thickness of 3.0 mm and in-plane resolution of 0.35 mm. The H&N PET scan was acquired on a GE Discovery 600 device with a slice thickness of 3.3 mm and in-plane resolution of 2.73 mm. The GI PET scan was acquired on a GE Discovery STE device with a slice thickness of 3.3 mm and in-plane resolution of 5.47 mm.

**Table 2.** Computed tomography (CT) acquisition parameters for cases included in this data descriptor. Cases included breast, sarcoma, head and neck (H&N), gynecologic (GYN), and gastrointestinal (GI) cancer.

| CT Acquisition Parameter | Breast | Sarcoma | H&N | GYN | GI |
| --- | --- | --- | --- | --- | --- |
| Manufacturer | GE | SIEMENS | SIEMENS | GE | GE |
| Model | Discovery CT590 RT | SOMATOM Confidence | Sensation Open | Discovery CT590 RT | Discovery CT590 RT |
| Slice Thickness (mm) | 2.5 | 3 | 3 | 2.5 | 2.5 |
| KVP (kV) | 120 | 120 | 120 | 120 | 120 |
| Exposure Time (ms) | 891 | 1000 | 1000 | 856 | 856 |
| X-Ray Tube Current (mA) | 154 | 111 | 32 | 167 | 277 |
| Rows | 512 | 512 | 512 | 512 | 512 |
| Columns | 512 | 512 | 512 | 512 | 512 |
| In-plane Resolution (mm) | 1.26 | 1.26 | 0.98 | 0.98 | 0.98 |
| Reconstruction diameter (mm) | 650 | 650 | 500 | 500 | 500 |
| Number of axial slices | 140 | 229 | 143 | 195 | 196 |

### IRB exemption and data storage

The retrospective acquisition, storage, and use of these DICOM files have been reviewed by the Memorial Sloan Kettering (MSK) Human Research Protection Program (HRPP) Office on 05/26/2021 and were determined to be exempt research as per 45 CFR 46.104(d)(3),(i)(a), (ii) and (iii), (i)(b),(ii) and (iii), (i)(c), (ii) and (iii) and 45.CFR.46.111(a)(7)). A limited IRB review of the protocol X19-040 A(1) was conducted via expedited process in accordance with 45 CFR 46.110(b), and the protocol was approved on May 26, 2021. DICOM files were obtained and stored on MIMcloud (MIM Software Inc., Ohio, USA), which is a HIPAA-compliant cloud-based storage for DICOM image files that has been approved for use at MSK by MSK’s Information Security team.

### DICOM anonymization

For each image, the DICOM header tags containing the patient name, date of birth, and patient identifier number were consistently removed from all DICOM files using DicomBrowser v. 1.5.2 ^20^. The removal of acquisition data and time metadata (if available in DICOM header tags) caused compatibility issues with ProKnow so were kept as is. Moreover, if institution name or provider name were available in the DICOM file, they were not removed as they were not considered protected health information. Select cases (breast, GYN, GI) were previously anonymized using the DICOM Import Export tool (Varian Medical Systems, CA, USA).

### Participant details

To register for the challenge, participants completed a baseline questionnaire that included their name, email address, affiliated institution, country, specialization, years in practice, number of disease sites treated, volume of patients treated per month for the designated tumor site, how they learned about this challenge, and reasons for participation. Registrant intake information was collected through the Research Electronic Data Capture (REDCap) system - a widely used web application for managing survey databases ^21^; an example of the intake form can be found at: https://redcap.mskcc.org/surveys/?s=98ARPWCMAT. The research conducted herein was approved by the HRRP at MSK (IRB#: X19-040 A(1); approval date: May 26, 2021). All subjects prospectively consented to participation in the present study, as well as to the collection, use, and disclosure of de-identified aggregate subject information and responses. Participants were categorized as recognized experts or non-experts. Recognized experts were identified by our C3RO team (EFG, CDF, DL) based on participation in the development of national guidelines or other extensive scholarly activities. Recognized experts were board-certified physicians with expertise in the specific disease site. Non-experts were any participants not categorized as an expert for that disease site. All non-experts had some knowledge of human anatomy, with the majority being composed of practicing radiation oncologists but also included resident physicians, radiation therapists, and medical physicists. Worthy of note, a participant could only be considered an expert for one disease site, but could have participated as a non-expert for other disease sites. Out of 1,026 registrants, 221 participated in generating segmentations, which were used for this dataset; due to the low participation rate, participants may represent a biased sample of registrants. Of note, participants could provide segmentations for multiple cases. Additional demographic characteristics of the participants can be found in Lin & Wahid et al. ^19^.

### ProKnow segmentation platform

Participants were given access to the C3RO workspace on ProKnow (Elekta AB, Stockholm, Sweden). ProKnow is a commercially available radiotherapy clinical workflow tool that allows for centralization of data in a secure web-based repository; the ProKnow system has been adopted by several large scale medical institutions and is used routinely in clinical and research environments. Anonymized CT DICOM images for each case were imported into the ProKnow system for participants to segment; anonymized MRI and PET images were also imported for select cases as available. Each case was attributed a short text prompt describing the patient presentation along with any additional information as needed. Participants were allowed to utilize common image manipulation (scrolling capabilities, zooming capabilities, window leveling, etc.) and segmentation (fill, erase, etc.) tools for generating their segmentations. No auto-segmentation capabilities were provided to the participants, i.e., all segmentations were manually generated. Notably, for the sarcoma case, an external mask of the patient’s body and a mask of the left femur was provided to participants. Screenshots of the ProKnow web interface platform for the various cases are shown in **Fig. 2**.

**Figure 2.**
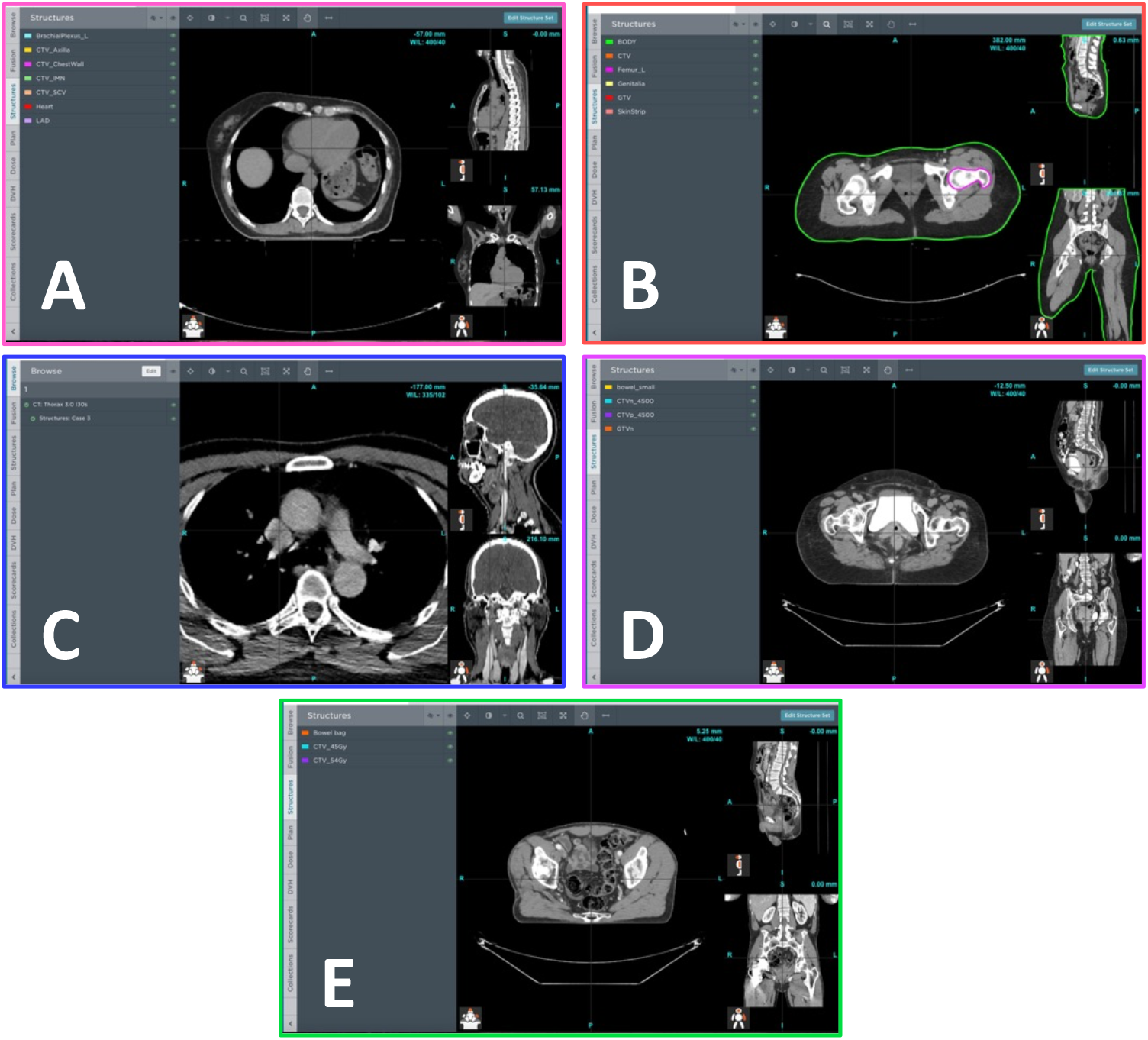
Examples of ProKnow segmentation platform used by participants for the breast **(A)**, sarcoma **(B)**, head and neck **(C)**, gynecologic **(D)**, and gastrointestinal **(E)** cases. Participants were given access to the standard image visualization and segmentation capabilities for generating their segmentations of target structures and organs at risk. Participants were also given access to a short prompt describing the patient presentation along with any additional information as needed. For the sarcoma case, an external mask of the patient’s body (green) and a mask of the left femur (pink) was provided to participants. Subplots for breast, sarcoma, head and neck, gynecologic, and gastrointestinal cases are outlined in pink, red, blue, purple, and green borders, respectively.

### Segmentation details

For each case, participants were requested to segment a select number of ROIs corresponding to target structures or OARs. Notably, not all participants generated segmentations for all ROIs. ROIs for each participant were combined into one structure set in the ProKnow system. ROIs were initially named in a consistent, but non-standardized format, so during file conversion ROIs were renamed based on The Report of American Association of Physicists in Medicine Task Group 263 (TG-263) suggested nomenclature ^22^; TG-263 was chosen due its ubiquity in standardized radiotherapy nomenclature. A list of each ROI and the number of available segmentations stratified by participant expertise level is shown in **Table 3**.

**Table 3.**
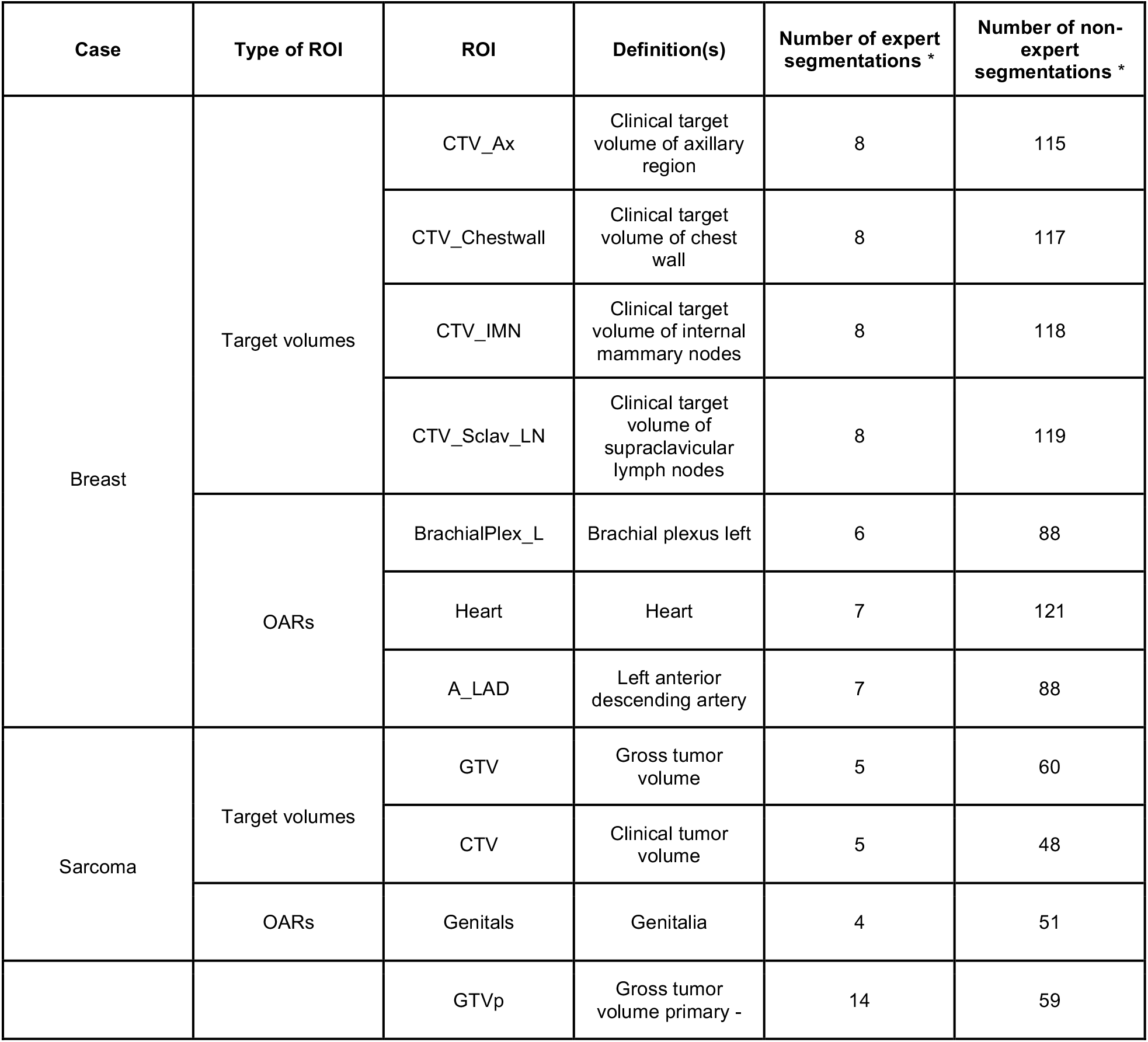
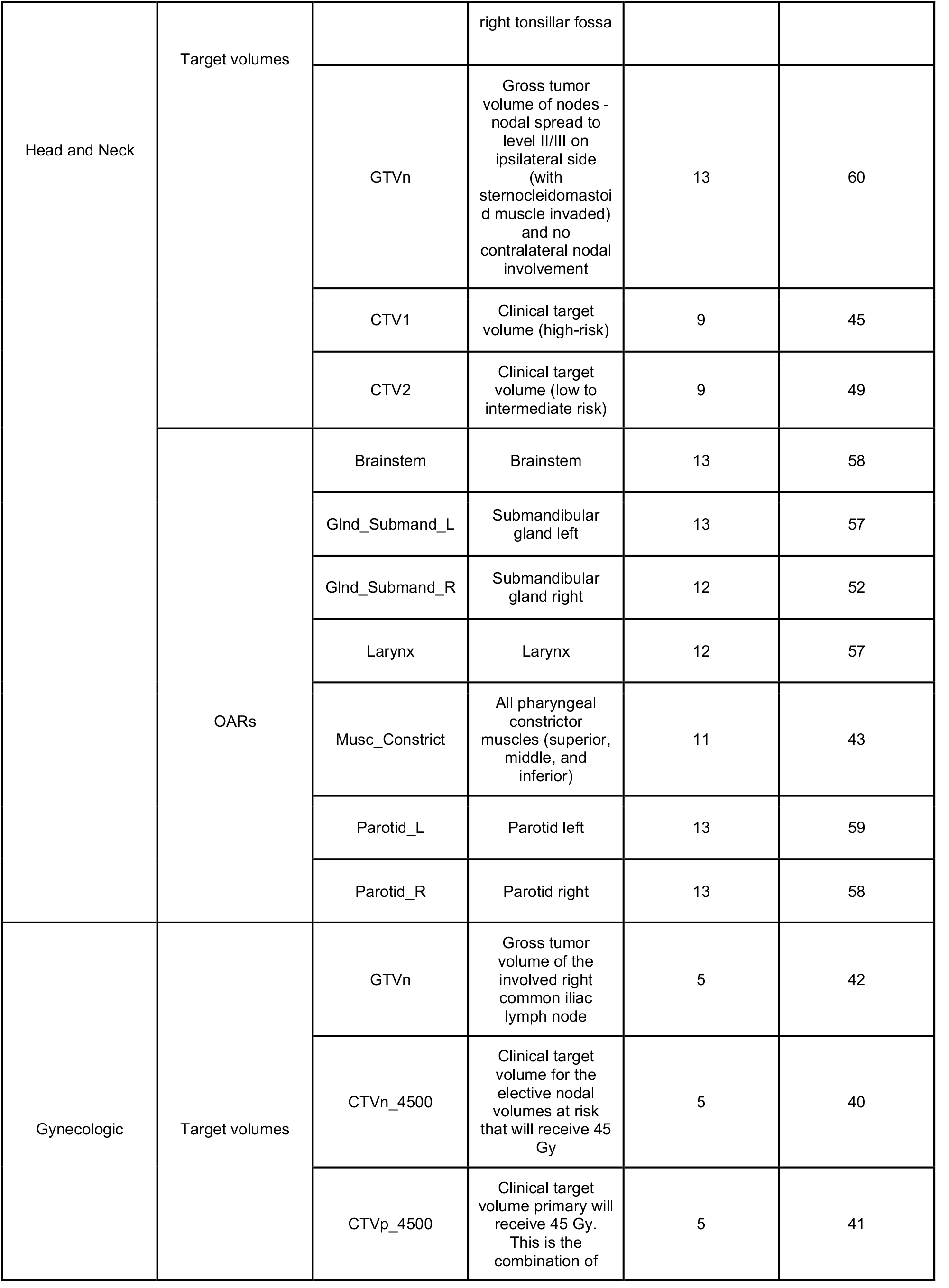
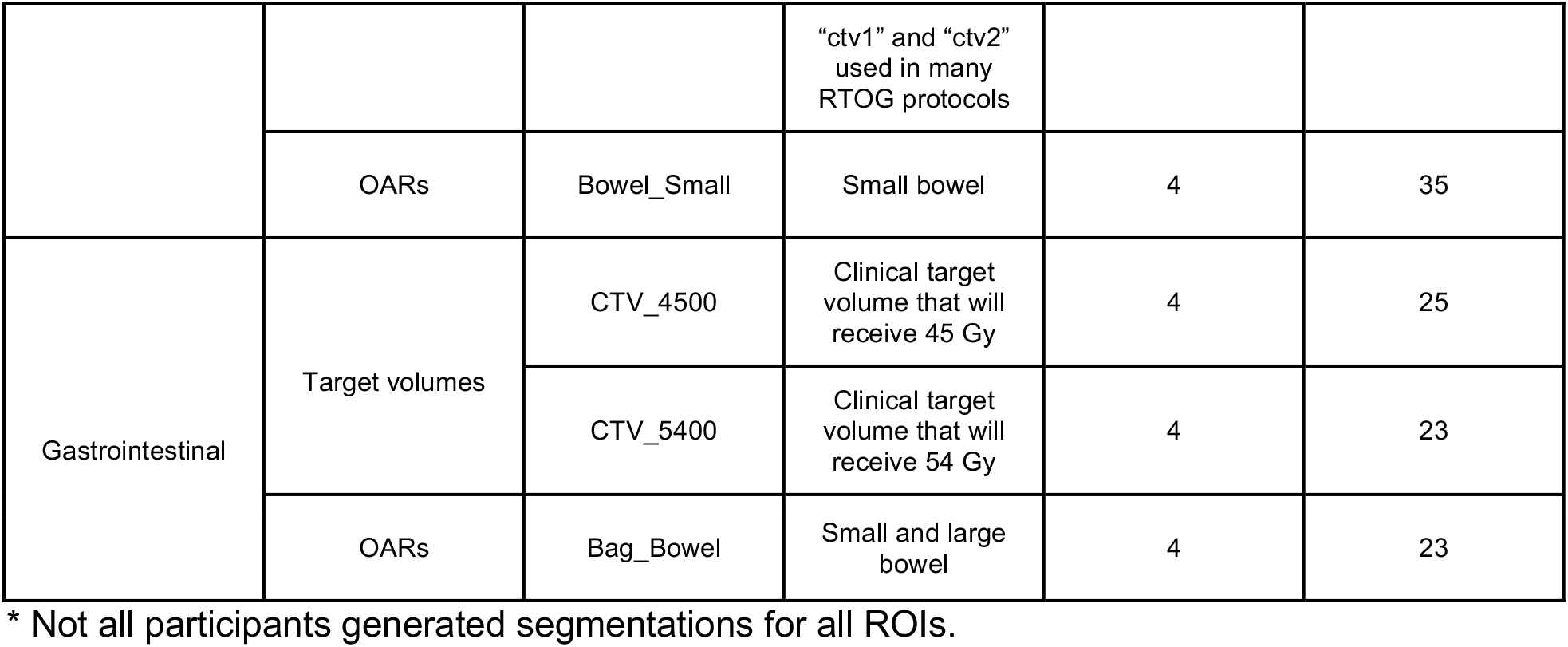
Summary of all region of interest (ROI) segmentations generated by participants for this data descriptor. ROIs included radiotherapy target volumes and organs at risk (OARs).

### Image processing and file conversion

For each case, anonymized CT images and structure sets for each annotator were manually exported from ProKnow in DICOM and DICOM radiotherapy structure (RTS) format, respectively. The Neuroimaging Informatics Technology Initiative (NIfTI) format is increasingly used for reproducible imaging research ^23–27^ due to its compact file size and ease of implementation in computational models ^28^. Therefore, in order to increase the interoperability of these data, we converted all our DICOM imaging and segmentation data to NIfTI format. For all file conversion processes, Python v. 3.8.8 ^29^ was used. An overview of the image processing workflow is shown in **Fig. 3A**. In brief, using an in-house Python script, DICOM images and structure sets were loaded into numpy array format using the DICOMRTTool v. 0.4.2 library ^30^, and then converted to NIfTI format using SimpleITK v. 2.1.1 ^31^. For each annotator, each individual structure contained in the structure set was separately converted into a binary mask (0 = background, 1 = ROI), and was then converted into separate NIfTI files. Notably, voxels fully inside and outside the contour are included and not include in the binary mask, respectively, while voxels that overlapped the segmentation (edge voxels) were counted as surface coordinates and included in the binary mask; additional details on array conversion can be found in the DICOMRTTool documentation ^30^. Examples of random subsets of five expert segmentations for each ROI from each case are shown in **Fig. 4**.

**Figure 3.**
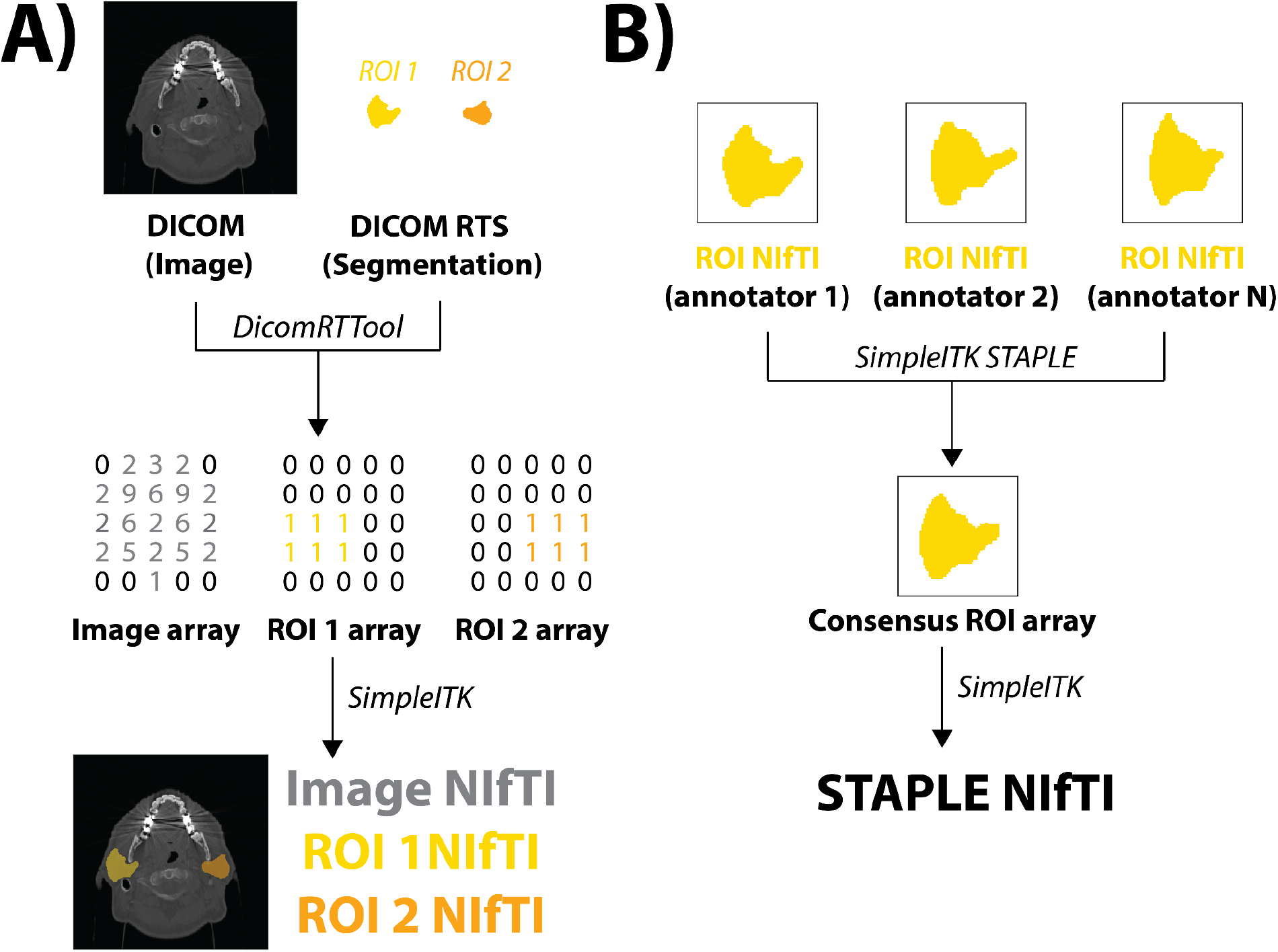
Image processing Python workflows implemented for this data descriptor for an example case (head and neck case, parotid glands). **(A)** Original Digital Imaging and Communications in Medicine (DICOM) formatted images and DICOM radiotherapy structure (RTS) formatted region of interest (ROI) segmentations are transformed to Neuroimaging Informatics Technology Initiative (NIfTI) format. **(B)**. ROIs from multiple annotators are combined into a single consensus segmentation using the Simultaneous Truth and Performance Level Estimation (STAPLE) method.

**Figure 4.**
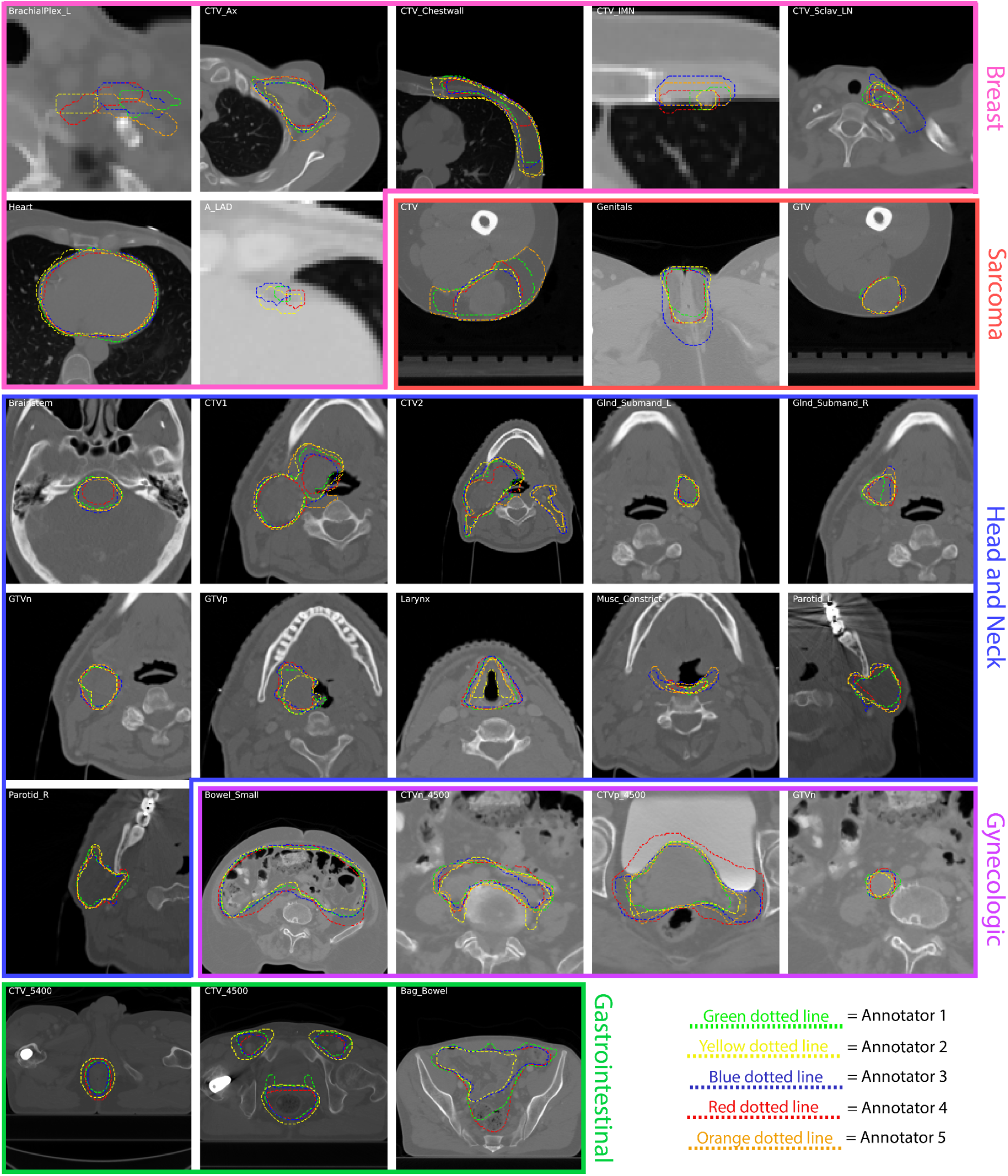
Examples of a random subset of five expert segmentations for each region of interest (ROI) provided in this data descriptor. Segmentations are displayed as green, yellow, blue, red, and orange dotted lines corresponding to annotators 1, 2, 3, 4, and 5, respectively, and overlaid on zoomed-in images for each case. Subplots for breast, sarcoma, head and neck, gynecologic, and gastrointestinal cases are outlined in pink, red, blue, purple, and green borders, respectively. Notably, the gastrointestinal case only had four expert annotators, so only four lines are displayed.

### Consensus segmentation generation

In addition to ground-truth expert and non-expert segmentations for all ROIs, we also generated consensus segmentations using the Simultaneous Truth and Performance Level Estimation (STAPLE) method, a commonly used probabilistic approach for combining multiple segmentations ^32–35^. Briefly, the STAPLE method uses an iterative expectation-maximization algorithm to compute a probabilistic estimate of the “true” segmentation by deducing an optimal combination of the input segmentations and incorporating a prior model for the spatial distribution of segmentations as well as implementing spatial homogeneity constraints ^36^. For our specific implementation of the STAPLE method, we utilized the SimpleITK STAPLE function with a default threshold value of 0.95. For each ROI, all available binary segmentation masks acted as inputs to the STAPLE function for each expertise level, subsequently generating binary STAPLE segmentation masks for each expertise level (i.e., STAPLE_expert_ and STAPLE_non-expert_). An overview of the consensus segmentation workflow is shown in **Fig. 3B**. Examples of STAPLE_expert_ and STAPLE_non-expert_ segmentations for each ROI are shown in **Fig. 5**.

**Figure 5.**
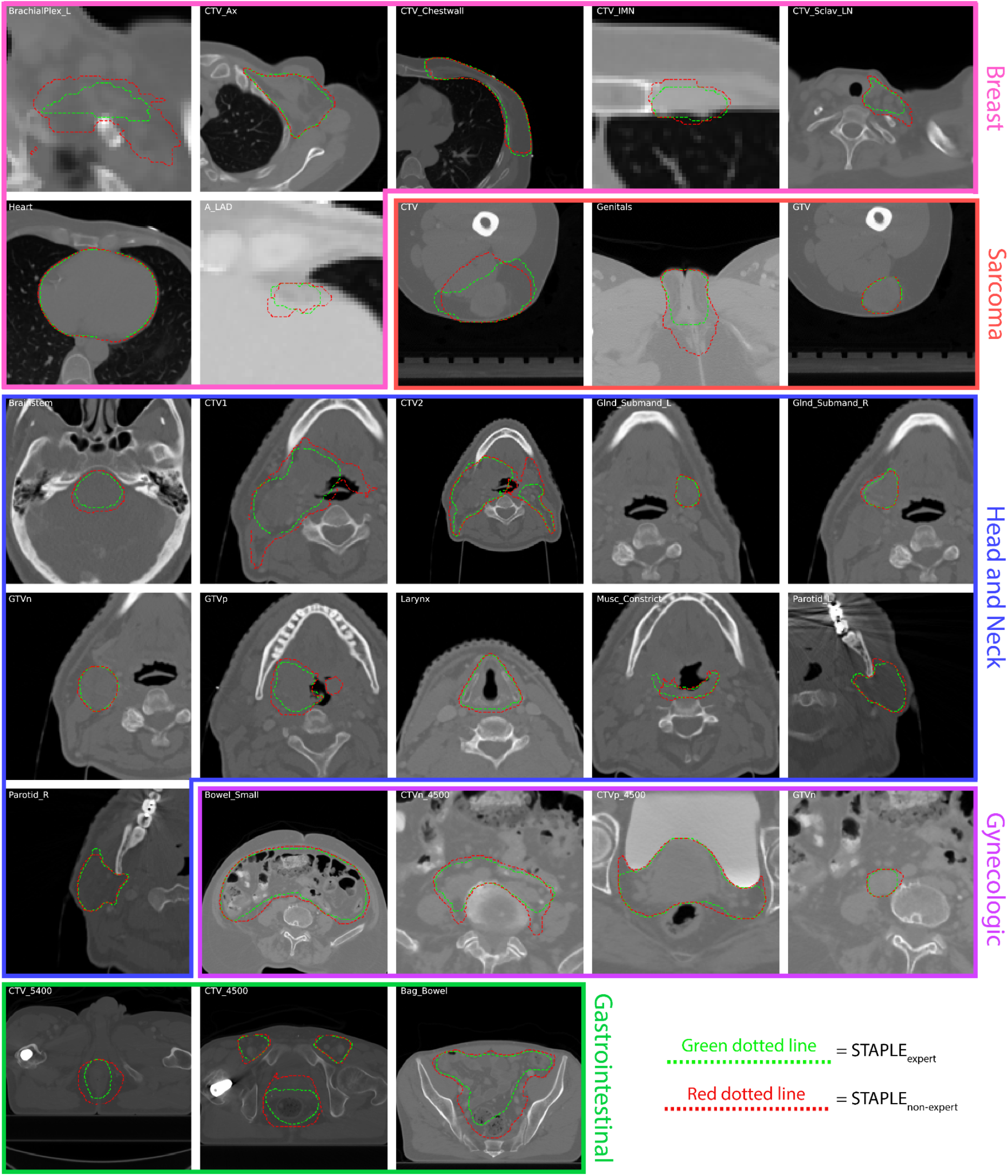
Examples of consensus segmentations using the simultaneous truth and performance level estimation (STAPLE) method for each region of interest (ROI) provided in this data descriptor. STAPLE segmentation generated by using all available expert segmentations (STAPLE_expert_) and STAPLE segmentation generated by using all available non-expert segmentations (STAPLE_non-expert_) are displayed as green and red dotted outlines, respectively, and overlaid on zoomed in images for each case. Subplots for breast, sarcoma, head and neck, gynecologic, and gastrointestinal cases are outlined in pink, red, blue, purple, and green borders, respectively.

## Data Records

### Medical images and multi-annotator segmentation data

This data collection primarily consists of 1985 3D volumetric compressed NIfTI files (.nii.gz file extension) corresponding to CT images and segmentations of ROIs from various disease sites (breast, sarcoma, H&N, GYN, GI). Analogously formatted MRI and PET images are available for select cases (sarcoma, H&N, GI). ROI segmentation NIfTI files are provided in binary mask format (0 = background,1 = ROI); file names for each ROI are provided in TG-263 notation. All medical images and ROI segmentations were derived from original DICOM and DICOM RTS files (.dcm file extension) respectively, which for completeness are also provided in this data collection. In addition, Python code to recreate the final NIfTI files from DICOM files is also provided in the corresponding GitHub repository (see *Code Availability* section).

### Consensus segmentation data

Consensus segmentations for experts and non-experts generated using the STAPLE method for each ROI have also been provided in compressed NIfTI file format (.nii.gz file extension). Consensus segmentation NIfTI files are provided in binary mask format (0 = background, 1 = ROI consensus). Python code to recreate the STAPLE NIfTI files from input annotator NIfTI files is also provided in the corresponding GitHub repository (see *Code Availability* section).

### Annotator demographics data

We also provide a single Microsoft Excel file (.xlsx file extension) containing each annotator’s gender, race/ethnicity, geographic setting, profession, years of experience, practice type, and categorized expertise level (expert, non-expert). Geographic setting was re-coded as “United States” or “International” to further de-identify the data. Each separate sheet corresponds to a separate disease site (sheet 1 = breast, sheet 2 = sarcoma, sheet 3 = H&N, sheet 4 = GU, sheet 5 = GI). Moreover, in order to foster secondary analysis of registrant data, we also include a sheet containing the combined intake data for all registrants of C3RO, including those who did not provide annotations (sheet 6).

### Folder structure and identifiers

Each disease site is represented by a top-level folder, containing a subfolder for images and segmentations. The annotator demographic excel file is located in the same top-level location as the disease site folders. Image folders contain separate subfolders for NIfTI format and DICOM format images. Segmentation folders contain separate subfolders for expert and non-expert segmentations. Each expertise folder contains separate subfolders for each annotator (which contains separate subfolders for DICOM and NIfTI formatted files) and the consensus segmentation (only available in NIfTI format). The data have been specifically structured such that for any object (i.e., an image or segmentation), DICOM and NIfTI subdirectories are available for facile partitioning of data file types. An overview of the organized data records for an example case is shown in **Fig. 6**. Segmentation files (DICOM and NIfTI) are organized by anonymized participant ID numbers and can be cross referenced against the excel data table using this identifier. The raw data, records, and supplemental descriptions of the meta-data files are cited under Figshare doi: 10.6084/m9.figshare.21074182 ^37^.

**Figure 6.**
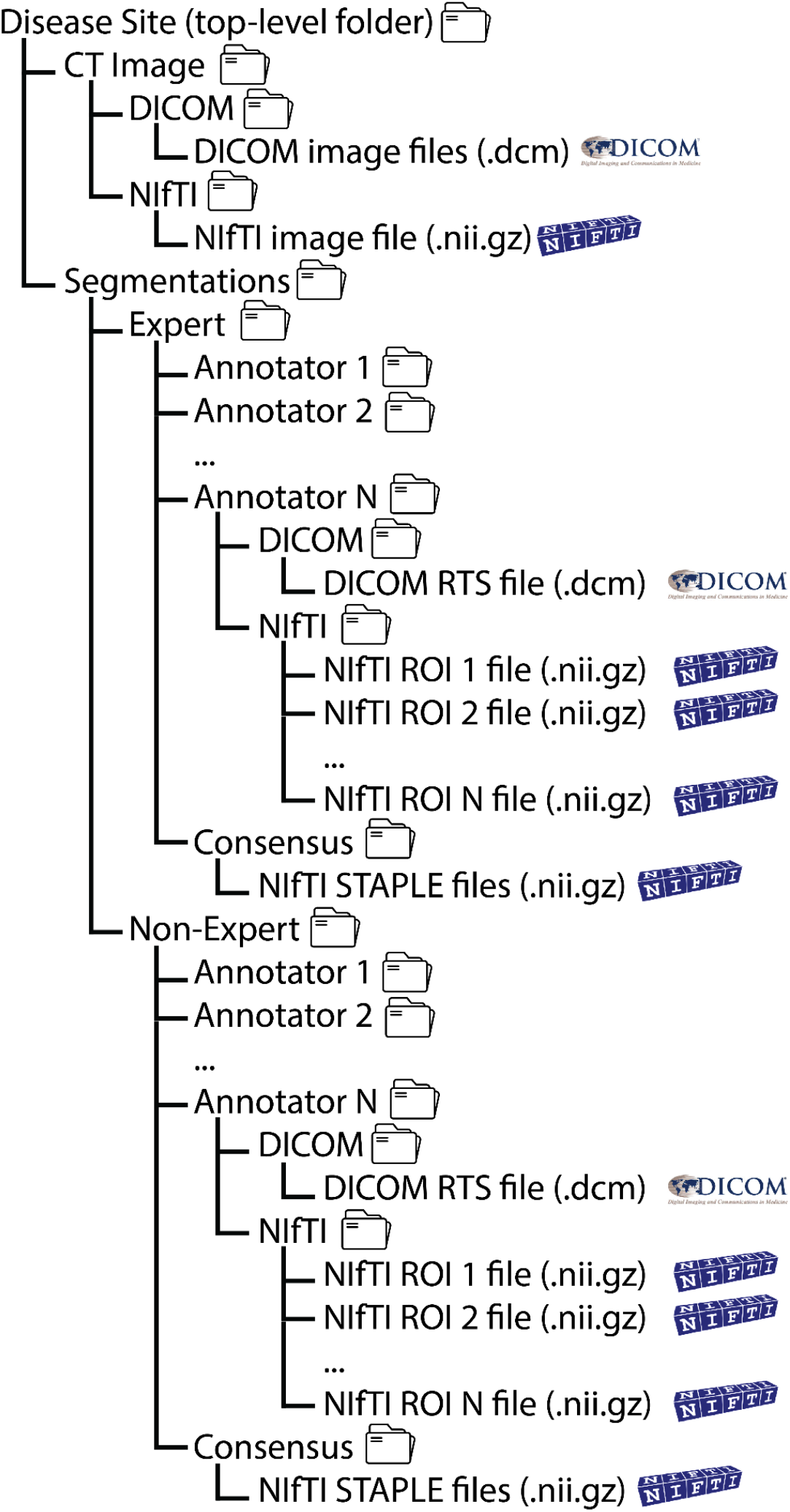
Overview of folder and file structure for dataset provided in this data descriptor. Each disease site folder contains separate subfolders for the computed tomography (CT) image and segmentations. Select cases had additional imaging modalities where available. Image subfolders contain separate subfolders for different data formats (Digital Imaging and Communications in Medicine [DICOM] and Neuroimaging Informatics Technology Initiative [NIfTI]). Segmentation subfolders contain separate subfolders which stratify expert and non-experts, which are further divided into subfolders for each annotator’s segmented ROIs in DICOM and NIfTI formats. Consensus segmentations for each ROI generated by the simultaneous truth and performance level estimation (STAPLE) method are also provided in expert and non-expert folders.

## Technical Validation

### Data annotations

Segmentation DICOM and NIfTI files were manually verified by study authors (D.L., K.A.W., O.S.) to be annotated with the appropriate corresponding ROI names.

### Segmentation interobserver variability

We calculated the pairwise interobserver variability (IOV) for each ROI for each disease site across experts and non-experts. Specifically, for each metric all pairwise combinations between all available segmentations in a given group (expert or non-expert) were calculated; median and interquartile range values are reported in **Table 4**.

**Table 4.** Pairwise interobserver variability values for experts and non-experts. Pairwise Dice similarity coefficient (DSC), average surface distance (ASD), and surface DSC (SDSC) are shown for experts and non-experts separately. Median values reported with interquartile range in parenthesis.

| Case | Type of ROI | ROI | Expert DSC | Expert ASD | Expert SDSC | Non-Expert DSC | Non-Expert ASD | Non-Expert SDSC |
| --- | --- | --- | --- | --- | --- | --- | --- | --- |
| Breast | Target volumes | CTV_Ax | 0.69<br>(0.07) | 3.41<br>(1.13) | 0.63<br>(0.11) | 0.61<br>(0.18) | 4.04<br>(2.89) | 0.60<br>(0.18) |
|  |  | CTV_Chestwall | 0.67<br>(0.14) | 4.44<br>(2.21) | 0.69<br>(0.16) | 0.68<br>(0.15) | 4.37<br>(2.47) | 0.68<br>(0.19) |
|  |  | CTV_IMN | 0.47<br>(0.14) | 2.71<br>(1.48) | 0.72<br>(0.16) | 0.36<br>(0.26) | 4.29<br>(5.09) | 0.59<br>(0.30) |
|  |  | CTV_Sclav_LN | 0.57<br>(0.12) | 3.65<br>(1.58) | 0.65<br>(0.14) | 0.56<br>(0.21) | 3.82<br>(2.65) | 0.63<br>(0.21) |
|  | OARs | BrachialPlex_L | 0.20<br>(0.27) | 4.19<br>(15.64) | 0.58<br>(0.52) | 0.25<br>(0.20) | 7.23<br>(9.53) | 0.59<br>(0.29) |
|  |  | Heart | 0.91<br>(0.08) | 1.84<br>(1.39) | 0.74<br>(0.13) | 0.93<br>(0.07) | 1.49<br>(1.18) | 0.75<br>(0.17) |
|  |  | A_LAD | 0.33<br>(0.13) | 4.59<br>(2.93) | 0.72<br>(0.12) | 0.31<br>(0.30) | 6.20<br>(8.85) | 0.62<br>(0.40) |
| Sarcoma | Target volumes | GTV | 0.94<br>(0.02) | 0.39<br>(0.24) | 0.80<br>(0.09) | 0.92<br>(0.14) | 0.47<br>(1.11) | 0.69<br>(0.40) |
|  |  | CTV | 0.72<br>(0.16) | 5.36<br>(4.66) | 0.72<br>(0.23) | 0.67<br>(0.31) | 4.73<br>(6.43) | 0.64<br>(0.47) |
|  | OARs | Genitals | 0.69<br>(0.04) | 3.19<br>(0.58) | 0.73<br>(0.06) | 0.58<br>(0.31) | 4.27<br>(5.93) | 0.61<br>(0.32) |
| Head and Neck | Target volumes | GTVp | 0.79<br>(0.06) | 1.40<br>(0.77) | 0.68<br>(0.15) | 0.74<br>(0.12) | 2.35<br>(3.44) | 0.62<br>(0.20) |
|  |  | GTVn | 0.91<br>(0.02) | 0.52<br>(0.16) | 0.64<br>(0.07) | 0.87<br>(0.10) | 0.84<br>(3.34) | 0.53<br>(0.20) |
|  |  | CTV1 | 0.85<br>(0.07) | 1.32<br>(0.77) | 0.65<br>(0.16) | 0.62<br>(0.34) | 6.75<br>(9.16) | 0.35<br>(0.32) |
|  |  | CTV2 | 0.71<br>(0.29) | 10.35<br>(12.58) | 0.83<br>(0.36) | 0.40<br>(0.45) | 18.98<br>(37.41) | 0.64<br>(0.43) |
|  | OARs | Brainstem | 0.82<br>(0.24) | 1.08<br>(1.20) | 0.74<br>(0.33) | 0.75<br>(0.16) | 1.59<br>(1.27) | 0.61<br>(0.26) |
|  |  | GInd_Submand_L | 0.86<br>(0.05) | 0.54<br>(0.24) | 0.65<br>(0.10) | 0.84<br>(0.15) | 0.65<br>(0.70) | 0.59<br>(0.24) |
|  |  | GInd_Submand_R | 0.80<br>(0.12) | 1.03<br>(0.95) | 0.78<br>(0.15) | 0.71<br>(0.30) | 1.27<br>(1.19) | 0.68<br>(0.23) |
|  |  | Larynx | 0.60<br>(0.30) | 2.17<br>(2.03) | 0.65<br>(0.33) | 0.60<br>(0.24) | 2.73<br>(1.69) | 0.54<br>(0.27) |
|  |  | Musc_Constrict | 0.58<br>(0.14) | 2.12<br>(1.14) | 0.76<br>(0.11) | 0.45<br>(0.24) | 2.84<br>(2.44) | 0.65<br>(0.27) |
|  |  | Parotid_L | 0.86<br>(0.04) | 0.90<br>(0.45) | 0.51<br>(0.08) | 0.80<br>(0.09) | 1.49<br>(1.23) | 0.40<br>(0.13) |
|  |  | Parotid_R | 0.87<br>(0.03) | 0.79<br>(0.34) | 0.51<br>(0.07) | 0.82<br>(0.10) | 1.31<br>(1.33) | 0.43<br>(0.14) |
| Gynecologic | Target volumes | GTVn | 0.79<br>(0.05) | 0.93<br>(3.19) | 0.43<br>(0.09) | 0.79<br>(0.45) | 1.28<br>(12.72) | 0.46<br>(0.40) |
|  |  | CTVn_4500 | 0.72<br>(0.03) | 3.03<br>(0.35) | 0.75<br>(0.06) | 0.66<br>(0.17) | 3.58<br>(2.12) | 0.68<br>(0.16) |
|  |  | CTVp_4500 | 0.79<br>(0.14) | 3.80<br>(2.48) | 0.75<br>(0.23) | 0.77<br>(0.13) | 3.63<br>(3.09) | 0.72<br>(0.20) |
|  | OARs | Bowel_Small | 0.80<br>(0.14) | 5.04<br>(3.02) | 0.70<br>(0.11) | 0.57<br>(0.40) | 8.38<br>(7.08) | 0.52<br>(0.27) |
| Gastrointestinal | Target volumes | CTV_4500 | 0.76<br>(0.03) | 4.10<br>(0.40) | 0.71<br>(0.08) | 0.65<br>(0.21) | 4.75<br>(3.12) | 0.66<br>(0.16) |
|  |  | CTV_5400 | 0.63<br>(0.20) | 15.22<br>(23.40) | 0.79<br>(0.40) | 0.48<br>(0.34) | 6.84<br>(5.90) | 0.87<br>(0.28) |
|  | OARs | Bag_Bowel | 0.64<br>(0.08) | 6.13<br>(2.50) | 0.65<br>(0.08) | 0.59<br>(0.27) | 7.68<br>(6.43) | 0.60<br>(0.21) |

Calculated metrics included the Dice Similarity coefficient (DSC), average surface distance (ASD), and surface DSC (SDSC). SDSC was calculated based on ROI specific thresholds determined by the median pairwise mean surface distance of all expert segmentations for that ROI as suggested in literature ^38^. Metrics were calculated using the Surface Distances Python package ^38,39^ and in-house Python code. For specific equations for metric calculations please see corresponding Surface Distances Python package documentation ^39^. Resultant values are broadly consistent with previous work in breast ^40^, sarcoma ^41^, H&N ^35,42,43^, GYN ^44^, and GI ^44–46^ IOV studies.

## Usage Notes

The image and segmentation data from this data collection are provided in original DICOM format (where applicable) and compressed NIfTI format with the accompanying excel file containing demographic information indexed by annotator identifiers. We invite all interested researchers to download this dataset for use in segmentation, radiotherapy, and crowdsourcing related research. Moreover, we encourage this dataset’s use for clinical decision support tool development. While the individual number of patient cases for this dataset is too small for traditional machine learning development (i.e., deep learning auto-segmentation training), this dataset could act as a benchmark reference for testing existing auto-segmentation algorithms. Importantly, this dataset could also be used as a standardized reference for future interobserver variability studies seeking to investigate further participant expertise criteria, e.g., true novice annotators (no previous segmentation or anatomy knowledge) could attempt to segment ROI structures on CT images, which could then be compared to our expert and non-expert annotators. Finally, in line with the goals of the eContour collaborative ^47^, these data could be used to help develop educational tools for radiation oncology clinical training.

The segmentations provided in this data descriptor have been utilized in a study by Lin & Wahid et al. ^19^. This study demonstrated several results that were consistent with existing literature, including: 1). target ROIs tended to exhibit greater variability than OAR ROIs ^35^, 2). H&N ROIs exhibited higher interobserver variability compared to other disease sites ^43,48^, and 3). non-expert consensus segmentations could approximate gold-standard expert segmentations ^49^.

Original DICOM format images and structure sets may be viewed and analyzed in radiation treatment planning software or select digital image viewing applications, depending on the end-user’s requirements. Current open-source software for these purposes includes ImageJ ^50^, dicompyler ^51^, ITK-Snap ^52^, and 3D Slicer ^53^ with the SlicerRT extension ^54^.

Processed NIfTI format images and segmentations may be viewed and analyzed in any NIfTI viewing application, depending on the end-user’s requirements. Current open-source software for these purposes includes ImageJ ^50^, ITK-Snap ^52^, and 3D Slicer ^53^.

## Data Availability

The raw data, records, and supplemental descriptions of the meta-data files are cited under Figshare doi: 10.6084/m9.figshare.21074182 (private until manuscript acceptance).

## Code Availability

Segmentations were performed using the commercially-available ProKnow (Elekta AB, Stockholm, Sweden) software. The code for NIfTI file conversion of DICOM CT images and corresponding DICOM RTS segmentations, along with code for consensus segmentation generation, was developed using in-house Python scripts and is made publicly available through GitHub: https://github.com/kwahid/C3RO_analysis.

## Acknowledgements

This work was supported by the National Institutes of Health (NIH)/National Cancer Institute (NCI) through a Cancer Center Support Grant (CCSG; P30CA016672-44; P30CA008748). D.L. is supported by the Radiological Society of North America (RSNA) Research Medical Student Grant (RMS2116). K.A.W. is supported by the Dr. John J. Kopchick Fellowship through The University of Texas MD Anderson UTHealth Graduate School of Biomedical Sciences, the American Legion Auxiliary Fellowship in Cancer Research, and an NIH/National Institute for Dental and Craniofacial Research (NIDCR) F31 fellowship (1 F31DE031502-01). E.F.G. and J.D.M. received funding from the Agency for Health Research and Quality (AHRQ R18HS026881). C.D.F. received funding from the NIH/NIDCR (1R01DE025248-01/R56DE025248); an NIH/NIDCR Academic-Industrial Partnership Award (R01DE028290); the National Science Foundation (NSF), Division of Mathematical Sciences, Joint NIH/NSF Initiative on Quantitative Approaches to Biomedical Big Data (QuBBD) Grant (NSF 1557679); the NIH Big Data to Knowledge (BD2K) Program of the NCI Early Stage Development of Technologies in Biomedical Computing, Informatics, and Big Data Science Award (1R01CA214825); the NCI Early Phase Clinical Trials in Imaging and Image-Guided Interventions Program (1R01CA218148); an NIH/NCI Pilot Research Program Award from the UT MD Anderson CCSG Radiation Oncology and Cancer Imaging Program (P30CA016672); an NIH/NCI Head and Neck Specialized Programs of Research Excellence (SPORE) Developmental Research Program Award (P50CA097007); and the National Institute of Biomedical Imaging and Bioengineering (NIBIB) Research Education Program (R25EB025787).

## Author Contributions Statement

Study conceptualization: C.D.F., E.F.G., B.E.N.; Study design: K.A.W., D.L., C.D.F., E.F.G., S.D., M.V.S., A.S.R.M., J.D.M.; Data acquisition: D.L., B.E.N., M.C., M.V.S., J.D.M.; Quality control of data and algorithms: K.A.W., D.L., O.S., R.H., M.A.N.; Manuscript editing: K.A.W., D.L., O.S., S.D., J.P.C, A.S.R.M. All authors contributed to the article and approved the submitted version.

## References

1. Sharp, G. et al. Vision 20/20: perspectives on automated image segmentation for radiotherapy. Med. Phys. 41, 050902 (2014).

2. Segedin, B. & Petric, P. Uncertainties in target volume delineation in radiotherapy – are they relevant and what can we do about them? Radiology and Oncology vol. 50 254–262 Preprint at https://doi.org/10.1515/raon-2016-0023 (2016).

3. Njeh, C. F. Tumor delineation: The weakest link in the search for accuracy in radiotherapy. J. Med. Phys. 33, 136–140 (2008).

4. Berry, S. L., Boczkowski, A., Ma, R., Mechalakos, J. & Hunt, M. Interobserver variability in radiation therapy plan output: Results of a single-institution study. Practical Radiation Oncology vol. 6 442–449 Preprint at https://doi.org/10.1016/j.prro.2016.04.005 (2016).

5. Sherer, M. V. et al. Metrics to evaluate the performance of auto-segmentation for radiation treatment planning: A critical review. Radiother. Oncol. 160, 185–191 (2021).

6. Cardenas, C. E., Yang, J., Anderson, B. M., Court, L. E. & Brock, K. B. Advances in Auto-Segmentation. Semin. Radiat. Oncol. 29, 185–197 (2019).

7. Harrison, K. et al. Machine Learning for Auto-Segmentation in Radiotherapy Planning. Clin. Oncol. 34, 74–88 (2022).

8. Yu, S. et al. Robustness study of noisy annotation in deep learning based medical image segmentation. Phys. Med. Biol. 65, 175007 (2020).

9. Budd, S. et al. Can Non-specialists Provide High Quality Gold Standard Labels in Challenging Modalities? in Domain Adaptation and Representation Transfer, and Affordable Healthcare and AI for Resource Diverse Global Health 251–262 (Springer International Publishing, 2021).

10. Heim, E. et al. Large-scale medical image annotation with crowd-powered algorithms. J Med Imaging (Bellingham) 5, 034002 (2018).

11. Wesemeyer, T., Jauer, M.-L. & Deserno, T. M. Annotation quality vs. quantity for deep-learned medical image segmentation. in Medical Imaging 2021: Imaging Informatics for Healthcare, Research, and Applications vol. 11601 63–76 (SPIE, 2021).

12. Afshar, P. et al. COVID-CT-MD, COVID-19 computed tomography scan dataset applicable in machine learning and deep learning. Sci Data 8, 121 (2021).

13. Wallner, J., Mischak, I. & Jan Egger. Computed tomography data collection of the complete human mandible and valid clinical ground truth models. Sci Data 6, 190003 (2019).

14. Lyu, X., Cheng, L. & Zhang, S. The RETA Benchmark for Retinal Vascular Tree Analysis. Sci Data 9, 397 (2022).

15. Kulaga-Yoskovitz, J. et al. Multi-contrast submillimetric 3 Tesla hippocampal subfield segmentation protocol and dataset. Sci Data 2, 150059 (2015).

16. Payette, K. et al. An automatic multi-tissue human fetal brain segmentation benchmark using the Fetal Tissue Annotation Dataset. Sci Data 8, 167 (2021).

17. Jin, K. et al. FIVES: A Fundus Image Dataset for Artificial Intelligence based Vessel Segmentation. Sci Data 9, 475 (2022).

18. Lin, D. et al. Contouring Collaborative for Consensus in Radiation Oncology (C3RO): An International Crowdsourcing Challenge to Improve Radiotherapy Contour Delineation. Int. J. Radiat. Oncol. Biol. Phys. 111, e10–e11 (2021).

19. Lin, D. et al. ‘E pluribus unum’: Prospective acceptability benchmarking from the contouring collaborative for consensus in Radiation Oncology (C3RO) crowdsourced initiative for multi-observer segmentation. medRxiv (2022) doi:10.1101/2022.09.23.22280295.

20. Archie, K. A. & Marcus, D. S. DicomBrowser: software for viewing and modifying DICOM metadata. J. Digit. Imaging 25, 635–645 (2012).

21. Harvey, L. A. REDCap: web-based software for all types of data storage and collection. Spinal Cord 56, 625 (2018).

22. Mayo, C. S. et al. American Association of Physicists in Medicine Task Group 263: Standardizing Nomenclatures in Radiation Oncology. Int. J. Radiat. Oncol. Biol. Phys. 100, 1057–1066 (2018).

23. Antonelli, M. et al. The Medical Segmentation Decathlon. Nat. Commun. 13, 4128 (2022).

24. Zbinden, L. et al. Convolutional neural network for automated segmentation of the liver and its vessels on non-contrast T1 vibe Dixon acquisitions. Sci. Rep. 12, 22059 (2022).

25. Chitalia, R. et al. Expert tumor annotations and radiomics for locally advanced breast cancer in DCE-MRI for ACRIN 6657/I-SPY1. Sci Data 9, 440 (2022).

26. Wahid, K. A. et al. Muscle and adipose tissue segmentations at the third cervical vertebral level in patients with head and neck cancer. Sci Data 9, 470 (2022).

27. Chiu, T.-W., Tsai, Y.-L. & Su, S.-F. Automatic detect lung node with deep learning in segmentation and imbalance data labeling. Sci. Rep. 11, 11174 (2021).

28. Li, X., Morgan, P. S., Ashburner, J., Smith, J. & Rorden, C. The first step for neuroimaging data analysis: DICOM to NIfTI conversion. J. Neurosci. Methods 264, 47–56 (2016).

29. Van Rossum, G. & Drake, F. L. Python 3 Reference Manual. (CreateSpace, 2009).

30. Anderson, B. M., Wahid, K. A. & Brock, K. K. Simple Python Module for Conversions Between DICOM Images and Radiation Therapy Structures, Masks, and Prediction Arrays. Pract. Radiat. Oncol. 11, 226–229 (2021).

31. Yaniv, Z., Lowekamp, B. C., Johnson, H. J. & Beare, R. SimpleITK Image-Analysis Notebooks: a Collaborative Environment for Education and Reproducible Research. J. Digit. Imaging 31, 290–303 (2018).

32. Taku, N. et al. Auto-detection and segmentation of involved lymph nodes in HPV-associated oropharyngeal cancer using a convolutional deep learning neural network. Clinical and Translational Radiation Oncology 36, 47–55 (2022).

33. Naser, M. A. et al. Head and Neck Cancer Primary Tumor Auto Segmentation Using Model Ensembling of Deep Learning in PET/CT Images. in Head and Neck Tumor Segmentation and Outcome Prediction 121–133 (Springer International Publishing, 2022).

34. McDonald, B. A. et al. Investigation of autosegmentation techniques on T2-weighted MRI for off-line dose reconstruction in MR-linac Adapt to Position workflow for head and neck cancers. bioRxiv (2021) doi:10.1101/2021.09.30.21264327.

35. Cardenas, C. E. et al. Comprehensive Quantitative Evaluation of Variability in Magnetic Resonance-Guided Delineation of Oropharyngeal Gross Tumor Volumes and High-Risk Clinical Target Volumes: An R-IDEAL Stage 0 Prospective Study. Int. J. Radiat. Oncol. Biol. Phys. 113, 426–436 (2022).

36. Warfield, S. K., Zou, K. H. & Wells, W. M. Simultaneous truth and performance level estimation (STAPLE): an algorithm for the validation of image segmentation. IEEE Trans. Med. Imaging 23, 903–921 (2004).

37. Contouring Collaborative for Consensus in Radiation Oncology. Large-scale crowdsourced radiotherapy segmentations across a variety of cancer sites. Figshare https://doi.org/10.6084/m9.figshare.21074182.v3 (2022).

38. Nikolov, S. et al. Clinically Applicable Segmentation of Head and Neck Anatomy for Radiotherapy: Deep Learning Algorithm Development and Validation Study. J. Med. Internet Res. 23, e26151 (2021).

39. Livne, M., Hughes, C., Hawkins, P., Deason, L. & Dudovitch, G. surface-distance: Library to compute surface distance based performance metrics for segmentation tasks. GitHub https://github.com/deepmind/surface-distance (2018).

40. Arculeo, S. et al. The emerging role of radiation therapists in the contouring of organs at risk in radiotherapy: analysis of inter-observer variability with radiation oncologists for the chest and upper abdomen. Ecancermedicalscience 14, 996 (2020).

41. Dionisio, F. C. F. et al. Manual versus semiautomatic segmentation of soft-tissue sarcomas on magnetic resonance imaging: evaluation of similarity and comparison of segmentation times. Radiol Bras 54, 155–164 (2021).

42. van der Veen, J., Gulyban, A. & Nuyts, S. Interobserver variability in delineation of target volumes in head and neck cancer. Radiother. Oncol. 137, 9–15 (2019).

43. van der Veen, J., Gulyban, A., Willems, S., Maes, F. & Nuyts, S. Interobserver variability in organ at risk delineation in head and neck cancer. Radiat. Oncol. 16, 120 (2021).

44. Aklan, B. et al. Regional deep hyperthermia: impact of observer variability in CT-based manual tissue segmentation on simulated temperature distribution. Phys. Med. Biol. 62, 4479–4495 (2017).

45. Rasing, M. J. A. et al. Online adaptive MR-guided radiotherapy: Conformity of contour adaptation for prostate cancer, rectal cancer and lymph node oligometastases among radiation therapists and radiation oncologists. Tech Innov Patient Support Radiat Oncol 23, 33–40 (2022).

46. Franco, P. et al. Variability of clinical target volume delineation for rectal cancer patients planned for neoadjuvant radiotherapy with the aid of the platform Anatom-e. Clin Transl Radiat Oncol 11, 33–39 (2018).

47. Sherer, M. V. et al. Development and Usage of eContour, a Novel, Three-Dimensional, Image-Based Web Site to Facilitate Access to Contouring Guidelines at the Point of Care. JCO Clin Cancer Inform 3, 1–9 (2019).

48. Brouwer, C. L. et al. 3D Variation in delineation of head and neck organs at risk. Radiat. Oncol. 7, 32 (2012).

49. O’Neil, A. Q., Murchison, J. T., van Beek, E. J. R. & Goatman, K. A. Crowdsourcing Labels for Pathological Patterns in CT Lung Scans: Can Non-experts Contribute Expert-Quality Ground Truth? in Intravascular Imaging and Computer Assisted Stenting, and Large-Scale Annotation of Biomedical Data and Expert Label Synthesis 96–105 (Springer International Publishing, 2017).

50. Abramoff, Magalhães & Ram. Image processing with ImageJ. Biophotonics int.

51. Panchal, A. & Keyes, R. SU-GG-T-260: dicompyler: an open source radiation therapy research platform with a plugin architecture. Med. Phys. 37, 3245–3245 (2010).

52. Yushkevich, P. A., Yang Gao & Gerig, G. ITK-SNAP: An interactive tool for semi-automatic segmentation of multi-modality biomedical images. Conf. Proc. IEEE Eng. Med. Biol. Soc. 2016, 3342–3345 (2016).

53. Pieper, S., Halle, M. & Kikinis, R. 3D Slicer. in 2004 2nd IEEE International Symposium on Biomedical Imaging: Nano to Macro (IEEE Cat No. 04EX821) 632–635 Vol. 1 (2004).

54. Pinter, C., Lasso, A., Wang, A., Jaffray, D. & Fichtinger, G. SlicerRT: radiation therapy research toolkit for 3D Slicer. Med. Phys. 39, 6332–6338 (2012).

